# Trabeculae microstructure parameters serve as effective predictors for marginal bone loss of dental implant in the mandible

**DOI:** 10.1101/2020.09.16.20195602

**Authors:** Hengguo Zhang, Jie Shan, Ping Zhang, Xin Chen, Hongbing Jiang

**Affiliations:** Jiangsu Key Laboratory of Oral Diseases, Nanjing Medical University, Nanjing 210029, Jiangsu Province, China; Department of Oral and Maxillofacial Surgery, The Affiliated Stomatological Hospital of Nanjing Medical University, Nanjing 210029, Jiangsu Province, China

**Keywords:** Dental Implant, Trabecular Bone, Marginal Bone Loss, Machine Learning

## Abstract

Marginal bone loss (MBL) is one of the leading causes of dental implant failure. This study aimed to investigate the feasibility of machine learning (ML) algorithms based on trabeculae microstructure parameters to predict the occurrence of severe MBL. Eighty-one patients (41 severe MBL cases and 40 normal controls) were involved in the current study. Four ML models, including support vector machine (SVM), artificial neural network (ANN), logistic regression (LR), and random forest (RF), were employed to predict severe MBL. The area under the receiver operating characteristic (ROC) curve (AUC), sensitivity, and specificity were used to evaluate the performance of these models. At the early stage of functional loading, severe MBL cases showed a significant increase of structure model index and trabecular pattern factor in peri-implant alveolar bone. The SVM model exhibited the best outcome in predicting MBL (AUC = 0.967, sensitivity = 91.67%, specificity=100.00%), followed by ANN (AUC = 0.928, sensitivity = 91.67%, specificity=93.33%), LR (AUC = 0.906, sensitivity = 91.67%, specificity = 93.33%), RF (AUC = 0.842, sensitivity = 75.00%, specificity = 86.67%). Together, ML algorithms based on the morphological variation of trabecular bone can be used to predict severe MBL.

## 1. Introduction

Peri-implant bone tissue is fundamental for the initial stability and long-term survival of dental implants, and marginal bone resorption around the implant could result in implant failure^1,2^. Currently, acceptable standards of implant success are defined as marginal bone loss (MBL) less than 1.5-2.0 mm after the first year of functional loading and subsequently less than 0.2 mm per year^3-5^. Although radiographic assessment of MBL has been considered as an authoritative criterion to evaluate implant success^3,6^, how to predict MBL remains unclear. Therefore, it is urgent to find an effective method to predict MBL and the survival rates of implants.

It has been widely acknowledged that MBL is a multicausal condition with different risk factors playing their roles simultaneously^7,8^. At the early post-loading stage, occlusal force transmits through the implant to the alveolar bone and promotes bone remolding. Plenty of morphology parameters can exhibit the morphological and mechanical properties of trabecular bone in this process^9-11^. Other factors that might impact bone remolding or MBL include the configuration of the dental implant^9,12^, cortical bone thickness^10^, periodontal disease susceptibility^13^, and smoking^14^. Some researchers have employed linear model to predict MBL based on bone structure parameters with sensitivity of 62.1% and specificity of 67.5%^15^. Hence, the establishment of a precise model to predict MBL is still a challenging exploration.

Machine learning (ML) can utilize statistical and optimization techniques to learn and detect in-depth relationships from complex and large data sets^16^. ML has already been applied in many aspects of the medical domain, including disease detection, diagnosis, and treatment^17-19^. Notably, prognosis prediction of dental implant which based on ML model has been applied in several clinical research^20,21^. These results prompt us to investigate whether ML models can predict MBL more accurately than conventional statistical methods.

This study aimed to find an effective way to predict the occurrence of MBL. We hypothesized that ML algorithms combined with clinical and CBCT data could predict MBL more accurately than conventional methods. Four ML models, including Support Vector Machine (SVM), Artificial Neural Network (ANN), Logistic Regression (LR), and Random Forest (RF), were employed to predict MBL. ML models showed superior performance compared to the single predictor in predicting MBL of mandibular implant.

## 2. Results

### 2.1. Trabecular microarchitecture changes in severe MBL cases

Eighty-one subjects were included in this study, and forty-one were identified severe MBL of dental implants. Gender (P = 0.007), cortical bone thickness (P = 0.007), and smoking (P = 0.008) showed a significant difference between the severe MBL cases and the normal controls (Supplementary Table S1). Morphological variables of the peri-implant and the normal adjacent alveolar bone were also compared between the severe MBL cases and the normal controls. Structure model index (SMI) (P = 0.007) and trabecular pattern factor (Tb.Pf) (P = 0.017) significantly increased in peri-implant alveolar bone of severe MBL cases (Fig. 1). Percent bone volume (BV/TV) (P = 0.002), trabecular number (Tb.N) (P = 0.025), and intersection surface (I.S) (P = 0.030) significantly increased in peri-implant alveolar bone of normal controls (Fig. 1). Additionally, we analyzed preoperative trabecular microarchitecture parameters of all subjects at T_0_, and there was no difference between the two groups. The results of trabecular microarchitecture variables at T_1_ and T_0_ was exhibited in Supplementary Table S2 and Supplementary Table S3.

**Fig. 1.**
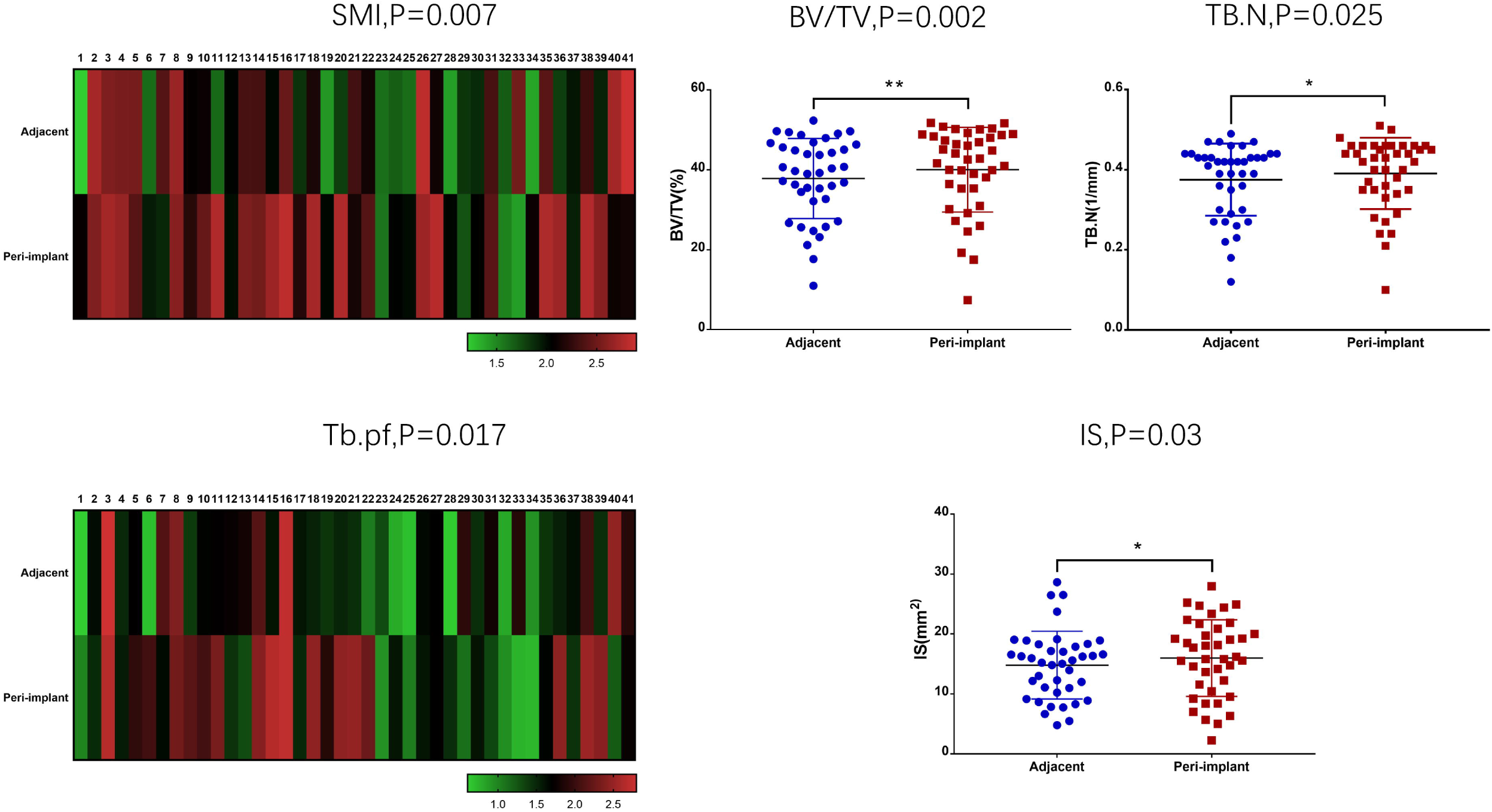
Comparison of morphological parameters among the peri-implant and normal adjacent alveolar bone in cases and controls. In severe MBL cases, SMI and Tb.Pf showed the visible difference between the peri-implant and normal adjacent alveolar bone. BV/TV, Tb.N, and i.S exhibited a significant difference between the peri-implant and normal adjacent alveolar bone in normal controls.

### 2.2. Morphological variables and their role in predicting MBL

Inspired by the above findings, we analyzed all variables using principal component analysis and correlation covariance matrices. All results relevant to morphological variables were confirmed with a significant difference and reasonable collinearity. SMI, Tb.Pf, Tb.N, bone surface volume ratio (BS/BV), and BV/TV manifested a higher correlation with MBL, while other morphological variables could not bring a noteworthy contribution. Fig. 2 reflected the ordination and contribution of all variables, along the first two “Multiple Factor Analysis” (MFA) components. The components explained 47.2% of the total variance in the data. Morphological parameters located in component 1 made significant contributions to the principal component, and all clinical parameters distributed in component 2.

**Fig. 2.**
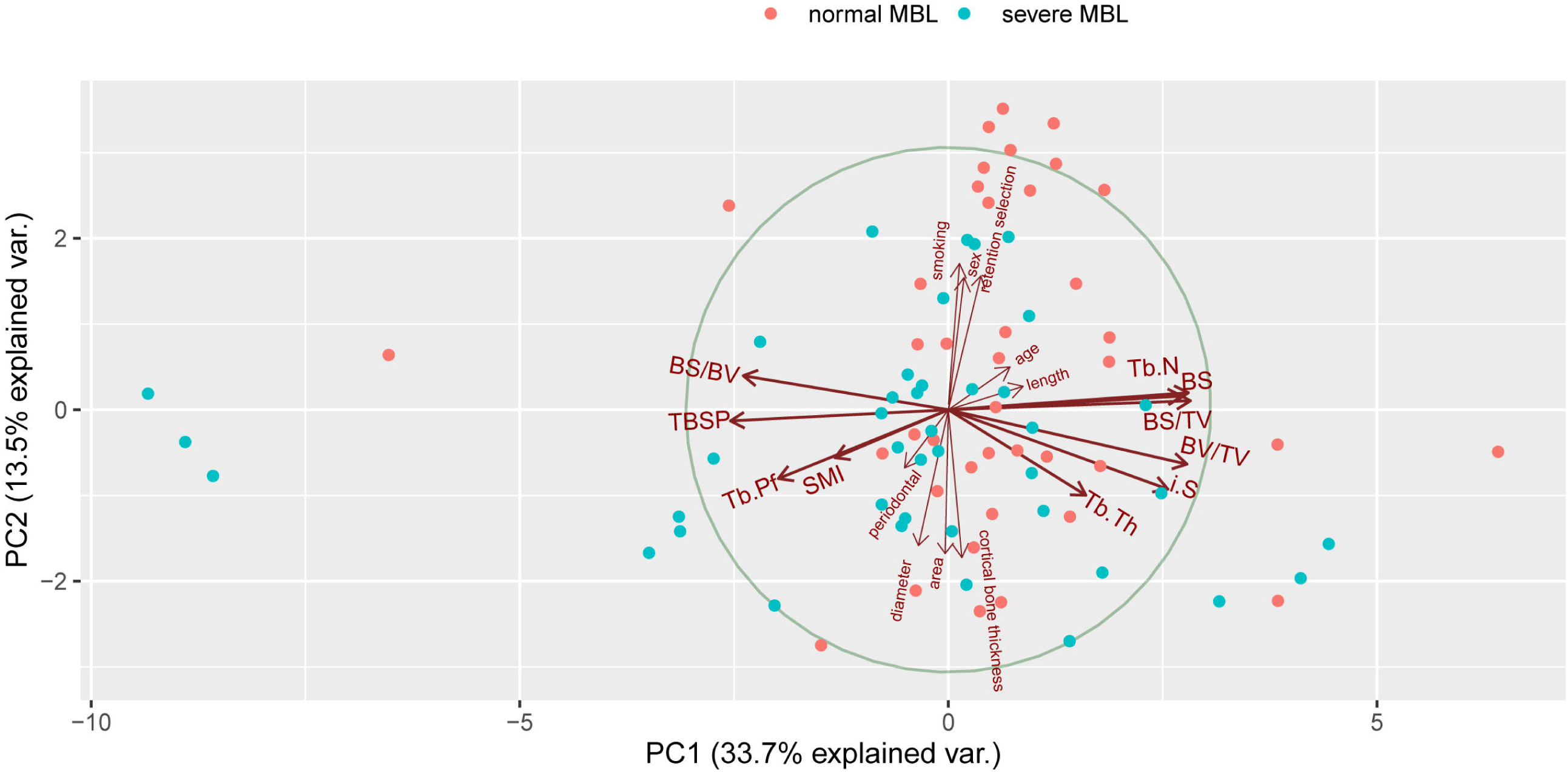
Plots of all variables in MFA. Closely clustered variables were positively correlated, while variables in opposing directions were negatively correlated. The length of the vector represented the importance of the variable in the MFA. Variables close to the midpoint of the circle plot had low contribution and weightage in the projection.

The logically obvious correlation between morphological parameters was shown in Fig. 3, and almost all correlation coefficients reached remarkably significant levels. Meanwhile, the linearity and credibility of trabecular bone microparameters were verified. SMI (P = 0.002) and Tb.Pf (P = 0.0165) exhibited a significantly high positive correlation with MBL. However, BV/TV and BS/BV manifested a negative correlation with MBL. Gender (P = 0.007), cortical bone thickness (P = 0.0072), and smoking (P = 0.0079) were powerfully correlated with MBL.

**Fig. 3.**
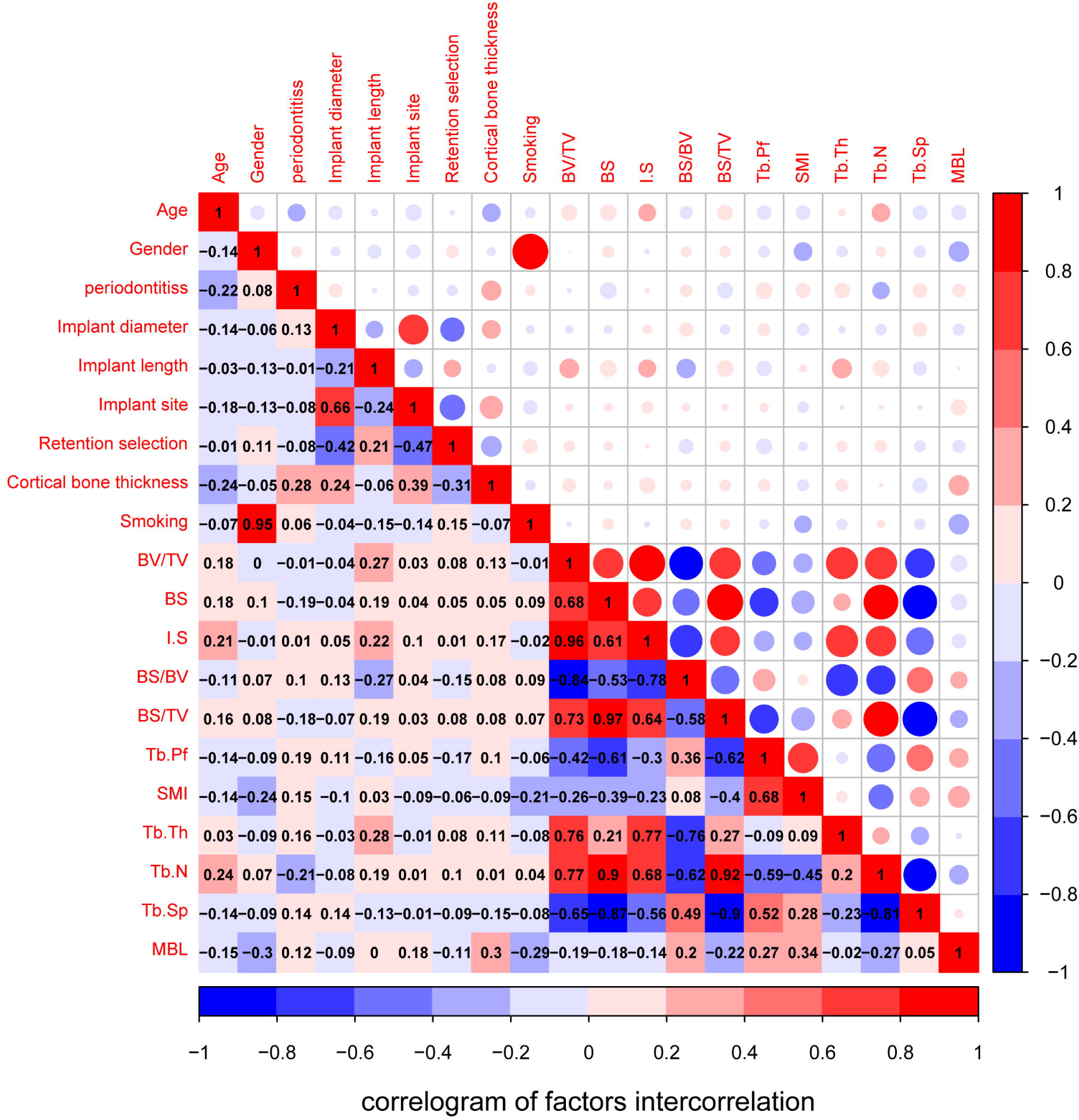
The visualization of correlation and covariance matrices between all variables. Red and blue represented positive and negative correlations, respectively. Darker colors indicated a more significant correlation.

### 2.3. Performance of ML models

Based on the consequence of correlation analysis, we eliminated some meaningless variables to build ML models. Each model was superior to a single factor predictor. The SVM model performed the best (AUC = 0.967), followed by ANN (AUC = 0.928), LR (AUC = 0.906), RF (AUC = 0.842), SMI alone (AUC = 0.705), Tb.Pf alone (AUC = 0.663), and BV/TV alone (AUC = 0.629) (Fig. 4, Table 1). As the best model, the SVM’s sensitivity and specificity were 91.67% and 100.00% at its optimal cutoff, respectively. SVM also presented a perfect optimal criterion (0.917), satisfied positive (1.000) and negative (0.938) predictive values, the maximum positive diagnose-likelihood-ratio (Infinite), and the minimum negative diagnose-likelihood-ratio (0.083). Moreover, the SVM model had the smallest false positivity and false negativity. The cutoff value of SMI was 1.027, while the corresponding sensitivity and specificity were 65.85% and 67.50%, respectively. The cutoff value of Tb.Pf was 0.968, and the sensitivity and specificity were 63.41% and 62.50%, respectively. We listed the performance of each model in Table 2. Based on the RF model, we rearranged the variables according to the importance of predicting MBL measurement through the Gini index (Fig. 5).

**Table 1.** Statistical significance of the difference between the areas under ROC curves. DeLong’s test and Bootstrap test were used.

|  | ANN | LR | RF | SMI alone | Tb.Pf alone | BV/TV alone |
| --- | --- | --- | --- | --- | --- | --- |
|  | *P*-value | *P*-value | *P*-value | *P*-value | *P*-value | *P*-value |
| Support vector machine (SVM) | 0.268 | 0.243 | 0.078 | P < 0.001*** | P < 0.001*** | P < 0.001*** |
| Artificial neural network(ANN) |  | 0.410 | 0.206 | 0.004** | P < 0.001*** | P < 0.001*** |
| Logistic regression (LR) |  |  | 0.199 | 0.023* | 0.009** | 0.003** |
| Random forest (RF) |  |  |  | 0.088 | 0.040* | 0.018* |
| SMI alone |  |  |  |  | 0.169 | 0.164 |
| Tb.Pf alone |  |  |  |  |  | 0.345 |

\* $p < 0.05$ , \*\* $p < 0.01$ , \*\*\* $p < 0.001$

**Table 2.** Performance of each model at optimal cutoff point.

| Model | Sensitivity | specificity | Optimal<br>cutoff of<br>probability | Positive<br>Predictive<br>Value | Negative<br>Predictive<br>Value | DLR<br>Positive | DLR<br>Negative | False<br>positive | False<br>negative | Optimal<br>criterion |
| --- | --- | --- | --- | --- | --- | --- | --- | --- | --- | --- |
| Support vector machine (SVM) | 91.67% | 100.00% | 0.547 | 1.000 | 0.938 | Infinite | 0.083 | 0 | 1 | 0.917 |
| Artificial neural network(ANN) | 91.67% | 93.33% | 0.998 | 0.917 | 0.933 | 13.750 | 0.089 | 1 | 1 | 0.917 |
| Logistic regression (LR) | 91.67% | 93.33% | 0.824 | 0.917 | 0.933 | 13.750 | 0.089 | 1 | 1 | 0.917 |
| Random forest (RF) | 75.00% | 86.67% | 0.560 | 0.818 | 0.813 | 5.625 | 0.288 | 2 | 3 | 0.750 |
| SMI alone | 65.90% | 67.50% | 1.027 | 0.675 | 0.659 | 2.026 | 0.506 | 13 | 14 | 0.659 |
| Tb.Pf alone | 63.40% | 62.50% | 0.968 | 0.634 | 0.625 | 1.691 | 0.585 | 15 | 15 | 0.625 |
| BV/TV alone | 39.00% | 45.00% | 1.049 | 0.421 | 0.419 | 0.710 | 1.355 | 22 | 25 | 0.390 |

The optimal cutoff was considered as the point maximizing the sum of sensitivity and specificity.

**Fig. 4.**
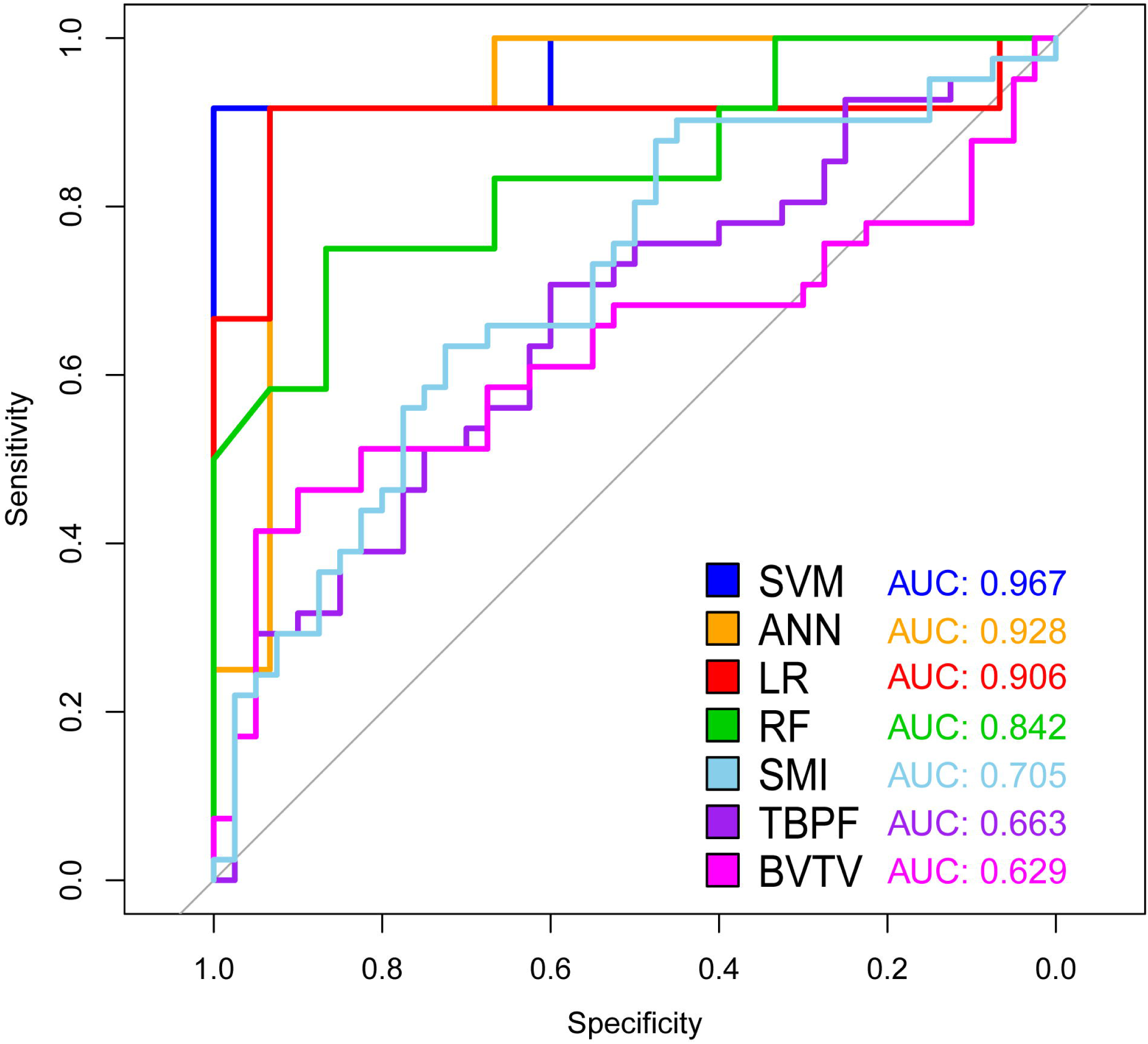
ROC & AUC of prediction models. The sensitivity and specificity of SVM, the best performing model, were 91.67% and 100.00%, respectively, at its optimal cutoff.

**Fig. 5.**
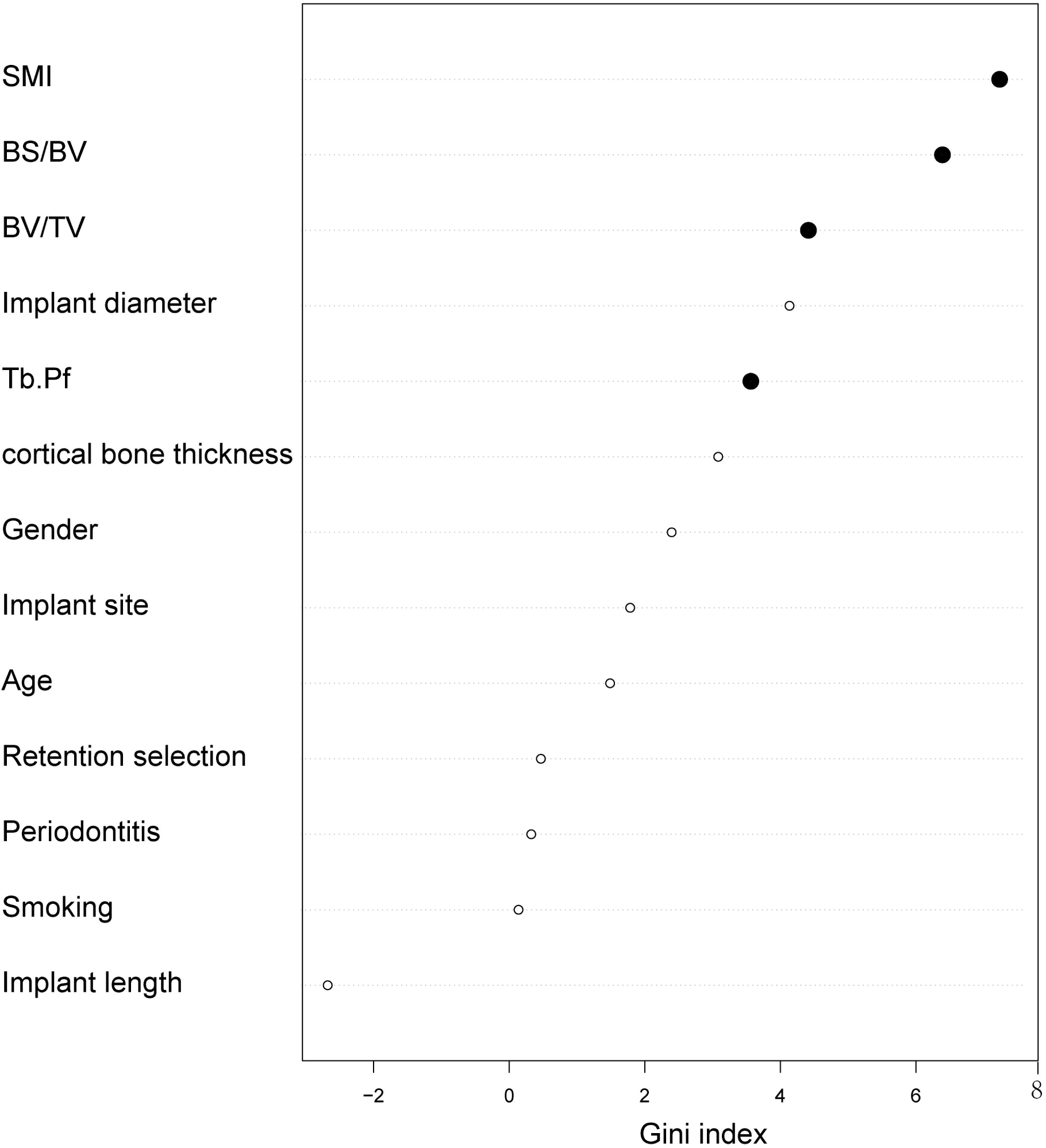
Variable importance plot of random forest model. The plot indicated the relative importance of the variables in the random forest model. Trabecular microarchitecture variables were marked as solid black points.

## 3. Discussion

Due to the important role of MBL in dental implant failure, MBL have become an essential clinical examination in postoperative follow-up. The purpose of this study was to verify that ML algorithms combined with early-stage trabecular bone variables could predict MBL more effectively than conventional methods. Our results showed a great performance of ML methods for predicting MBL, which can be considered as the feasible early warning for severe MBL. It was noted that the other factors resulting in MBL should be taken into account in future studies.

Previous researches about MBL mainly concentrated on the cause and treatment^6,9,12,13,22,23^, but rarely on the prediction of MBL^15^. Factors such as trabecular bone microstructure parameters possibly affecting MBL during bone remolding have been well elucidated^15^, but the role of trabecular bone in this progression remains unclear. To our knowledge, this is the first study to establish and validate ML models based on trabecular microstructure parameters to predict the occurrence of MBL of the submerged dental implant in mandible.

It has been widely acknowledged that various factors, including cortical bone thickness, smoking, periodontitis, SMI, Tb.Pf, and BV/TV, function as a complex to cause MBL^8^. Previous studies have also demonstrated that the proportion of cancellous bone^10^, crown-to-implant ratio^24^, bone texture, and cortical width^15^, are risk factors of MBL. Therfore, single predictive factor cannot accurately predict MBL because MBL is a multifactorial outcome. One recent study attempted to employ Cox regression and mixed linear modeling to predict the occurrence of MBL, but they aimed to assess interventions and their consequences with regard to further bone loss at sites diagnosed with peri-implant inflammation^25^. Another study incorporated several radiographic features of cortical and cancellous bone texture, cortical width, and patient smoking habits to build a statistical model to predict MBL with a sensitivity of 62.1% and specificity of 67.5%^15^. Compared to conventional statistical methods, the current study verified that ML algorithms predicted MBL more accurately. Of note, the SVM model performed best with a sensitivity of 91.67% and specificity of 100.00%, which was significantly better than that of SMI, Tb.Pf or BV/TV alone.

To demonstrate the differences and correlations of morphological variables between the controls and severe MBL cases, we also analyzed morphological variables of trabecular bone in patients with MBL during bone remolding. At the early stage of functional loading, CBCT analysis exhibited a worse outcome of SMI and Tb.Pf in peri-implant alveolar bone of severe MBL cases. These findings revealed that severe MBL cases demonstrated the premonitory morphological variation in trabecular microarchitecture at the early stage. Consistent with our results, a previous study reported that the preservation and improvement of trabecular microarchitecture always brought about a better therapeutic benefit for osteoporosis at multiple skeletal sites^26^. SMI and Tb.Pf were the best determinants of the MBL level, which they reflected the structure quality of the trabecular bone in the peri-implant of severe MBL cases. However, in normal controls, Tb.N and BV/TV were superior to other morphological variables ^11^.

Although the current study has demonstrated that ML could predict the occurrence of MBL more effectively than conventional methods, some limitations need to be acknowledged. The variables in the current study lacked results of the periodontal examination, such as probing depth. Additionally, this study was limited to patients who received implant treatment in mandible. Further studies on larger sample sizes using more relative variables (e.g. periodontal or microbiological) might be better for the ML performance. At last, a mean 20.95 ± 2.67 months of follow-up after functional loading was also limited to predict MBL. Hence, we entitled this study as a preliminary one.

In conclusion, the current study verified that the severity of MBL was closely related to trabecular microarchitecture during the early stage of functional loading. Change of trabecular microarchitecture can provide an early warning for severe MBL. ML models SVM, ANN, LR, and RF indicate superior performance compared to the single predictor in predicting MBL of mandibular implant.

## 4. Methods

### 4.1. Study design

To address the research purpose, we designed and implemented a cross-sectional study. All subjects receiving implant treatment between January 2016 and March 2019 in the Department of Oral and Maxillofacial Surgery of Affiliated Stomatological Hospital of Nanjing Medical University were screened. The inclusion criteria of subjects were as follows: 1) above 18 years of age with good health; 2) having received fixed prosthesis of the mandibular implant with at least one year of loading; 3) available integrated clinical data; 4) valid cone-beam computed tomography (CBCT) examination conducted at the following three time points: (T_0_) preoperative bone assessment, (T_1_) at the follow-up between three months and six months after loading, and (T_2_) at the follow-up above one year of post-loading. The exclusion criteria of subjects were as follows: 1) patients diagnosed with clinical or absolute failure based on the current guideline^4^; 2) patients with an incomplete periodontal follow-up examination and treatment record. 3) patients receiving bone augmentation. Patients with smoking cessation for more than three months before surgical implant placement were taken as non-smokers.

This study was approved by the ethics committee of the Affiliated Stomatological Hospital of Nanjing Medical University (Approval number: PJ2019-038-001, Approval data: March 15, 2019) in accordance with the Helsinki Declaration II. Written informed consents were obtained from all participants.

### 4.2. Measurement of MBL

All subjects were examined by the CBCT (NewTom 5G cone-beam computed tomography device, QR s.r.l, Verona, Italy) at a reconstruct voxel size of 150 μm at the three time points. Scanning voltage and current were 110 kV and 10 mA, while exposure and scanning times were 3.6 s and 18 s. We reconstructed the original radiographs using the center of the implant in the sagittal, coronal, and transverse plane. The implant length and diameter were used to test the accuracy of reconstructed images. MBL measurement was performed as follows: 1) the horizontal interface between implant and abutment was validated as the reference loci; 2) vertical distances from the loci to the most coronal level of bone to implant contact at the mesial and distal sites were measured at the preceding time points^27,28^; 3) analysis of radiographs was conducted by two investigators who did not participate in this study. We obtained the maximum MBL of the implant at T_1_ and T_2_ as the corresponding MBL level (Supplementary Figure S1). According to the T_2_ MBL level, we divided all subjects into two groups:

- Normal controls: less than 2 mm MBL in the first year after fixed prosthesis, then less than 0.2 mm MBL per year
- Severe MBL cases: MBL level exceeding normal controls

### 4.3. Measurement of peri-implant bone morphological parameters

We imported the T_1_ radiographs to CT Analyzer (CTAn, SkyScan, Antwerpen, Belgium). The threshold value of binary selection was determined by completely distinguish cortical bone and trabecular bone (Supplementary Figure S2a). We confirmed the region of interest (ROI) by the diameter of each implant. We selected five sequential ROI layers adjoining the implant root as the volume of interest (VOI) of peri-implant alveolar bone (Supplementary Figure S2b). Another five sequential ROI layers away from the implant were chosen as VOI of the normal adjacent alveolar bone (Supplementary Figure S2b). The trabecular bone morphological parameters, such as SMI, Tb.Pf, BV/TV, i.S, and Tb.N were extracted using three-dimensional analysis of each VOI. SMI, Tb.Pf, and i.S represent the shape and quality of trabecular bone. BV/TV and Tb.N usually mean quantity of trabecular bone. Finally, we calculated morphological variables by the ratio of peri-implant to normal adjacent alveolar bone.

### 4.4. PCA analysis

Including clinical and morphological parameters, all variables were utilized for ordination analysis and contribution degree evaluation of principle component using the “Multiple Factor Analysis” (MFA) function in the R package “FactoMineR”. As a dimensionality reduction method, MFA reduces the complexity of multivariate data and allows visual interpretation of significant patterns. It is suited to data that contains both continuous and categorical variables. MFA also allows grouping of variables where each group is normalized individually to balance their influence. An MFA correlation circle plot depicts the continuous variables, and the factors plot depicts the categorical variables.

### 4.5. Visualization of correlation and covariance matrices

Correlation and covariance matrices can visualize the patterns and relationships between the variables. We had twenty original variables, including object variable MBL. The visualization of matrices re-ordered the variables in a correlation matrix and displayed the value by sign and magnitude. All iconic encodings in the matrix displayed the pattern and significance level of correlations between variables. The R package “corrgram” and “Hmisc” were employed in this study.

### 4.6. ML algorithms

Based on the R Programming Language (R Core Team, Vienna, Austria), four ML models, including SVM, ANN, LR model and RF, were constructed. The dataset was randomly split into two mutually exclusive sets, training (70%) and testing (30%), a method called holdout method^29^. LR model established with variable choice through backward elimination was implemented to assess risk factors and predict the diagnosis of diseases. The R package “e1071” was applied in the SVM model to accomplish regression and classification missions by constructing hyperplanes in a multidimensional space. SVM model could manage multiple continuous and categorical variables according to the decision plane. ANN model, a computerized encoding of artificial humanoid neuronal networks, included the input layer, hidden layers, and output layer. Neurons connected the adjacent layers as a medium for the delivery-feedback-correction-delivery cycle. This recursive process adjusted the weights for fewer errors and better accuracy. ANN model was implemented by R package “neural net”. RF model, a ML algorithm based on the decision tree, could combine the output of a single decision tree to improve the overall performance. RF model was superior to a single decision tree in eliminating overfitting. RF model also could display the relative importance of the variables by the Gini index. We utilized the R package “randomForest” in the establishment of the RF model.

### 4.7. Statistical analysis

The chi-square test and Fisher’s exact test were applied to compare the variables of severe MBL cases and normal controls. We utilized the Cochran-Armitage trend test for categorical variables, while continuous variables were assessed by Student’s t-test and the Mann-Whitney rank-sum test. R programming language was used for all statistical analyses, while p < 0.05 was regarded as statistically significant.

## Data Availability

All CBCT files of patients and control subjects were stored in a non-public medical record database. CBCT data of the samples will not be shared.

## Acknowledgments

We would like to thank our colleagues Shen Xin and Lin Jialing from Jiangsu Key Laboratory of Oral Diseases, Nanjing Medical University, for their support of radiographs analysis in this project.

## Author contributions

The study was designed by HZ and HJ. The data was collected by HZ and analyzed by JS. The ML models were built by HZ, JS and XC. The paper was written by HZ and revised by JS, PZ, and HJ. They all made great contributions to this study and should be listed as authors. All authors read and approved the manuscript.

## Figure Legends

Figure S1. Reference lines for measurement of marginal bone loss around dental implants. R = reference line; X = implant diameter; L = implant length; M = mesial measurement; D = distal measurement.

Figure S2. Measurement of peri-implant bone morphological parameters. (a) Radiograph grayscale was selected by the principle of showing trabecular bone (marked in red) completely. (b) Five sequential ROI layers adjoining the implant were selected as VOI of the peri-implant alveolar bone (marked in red). Five sequential ROI layers away from the implant were chosen as VOI of the normal adjacent alveolar bone (marked in blue).

